# Central diabetes insipidus induced by temozolomide: A literature Review

**DOI:** 10.1101/2021.02.18.21252028

**Authors:** Mohammad D. Hossain, Abu Bakar Siddik, Susmita Dey Pinky, Tonazzina Hossain Sauda, Fahtiha Nasreen, Pallab Sarker, Masum Rahman

**Affiliations:** Jalalabad Ragib Rabeya Medical College and hospital, Sylhet, Bangladesh; Department of Pain Medicine, Mayo Clinic, Jacksonville, Florida, USA; Chittagong Medical College and hospital, Panchlaish, Chittagong; Bangladesh Medical College and Hospital, Dhaka, Bangladesh; Armed Forces Medical College and hospital, Dhaka, Bangladesh; Sher E Bangla Medical College Barisal and hospital, Bangladesh; Department of Neurological Surgery, Mayo Clinic, Rochester, Minnesota, USA

**Keywords:** hypernatremia, Central Diabetes Insipidus, Diabetes Insipidus, temozolomide, CNS tumor

## Abstract

Temozolomide has been the most used chemotherapeutic drug for glioblastoma and various CNS malignancies. Although myelosuppression has the most severe adverse effect, central diabetes insipidus (CDI) has been found as an infrequent side effect. CDI is characterized by decreased antidiuretic hormone secretion from posterior pituitary, thereby the inability to concentrate the urine with variable degrees of polyuria and compensatory polydipsia. Following a comprehensive literature search of several databases from 1990 to October 2020, which were limited to the English language, patient data were analyzed to demonstrate the risk factors, severity, reversibility of the disease, and overall survival. Total nine cases found who developed CDI following TMZ treatment. All patients manifest hyperosmolar symptoms like polyuria and polydipsia within 3 to 12 weeks following temozolomide initiation. Clinical and laboratory features, therapeutic response to exogenous desmopressin, and clinical course have been summarized.

## Introduction

Central diabetes insipidus (CDI) is a rare condition defined by decreased secretion of antidiuretic hormone (ADH) from supraoptic nucleus, resulting in a urine concentrating defect. It manifests as polyuria, bothersome nocturia, and compensatory polydipsia for urinary water losses. An increased urinary free water loss leads to an elevated or high normal range-serum sodium and serum osmolarity that triggers thirst (figure-1)(1). Clinical manifestations result from hyperosmotic or hypernatremia and largely neurologic, characterized by lethargy, fatigue, irritability, confusion, and even seizure. Regardless, the severity of hypernatremia is related to the thirst sensation’s functionality and is relatively worse when neurologically impaired or in a situation of restricted drinkable water access. The pathognomic factors leading to central DI includes idiopathic cause, tumors, infiltrative diseases, surgery, trauma, and medications. Hereditary forms have also been reported (2–4).

**Figure-1:**
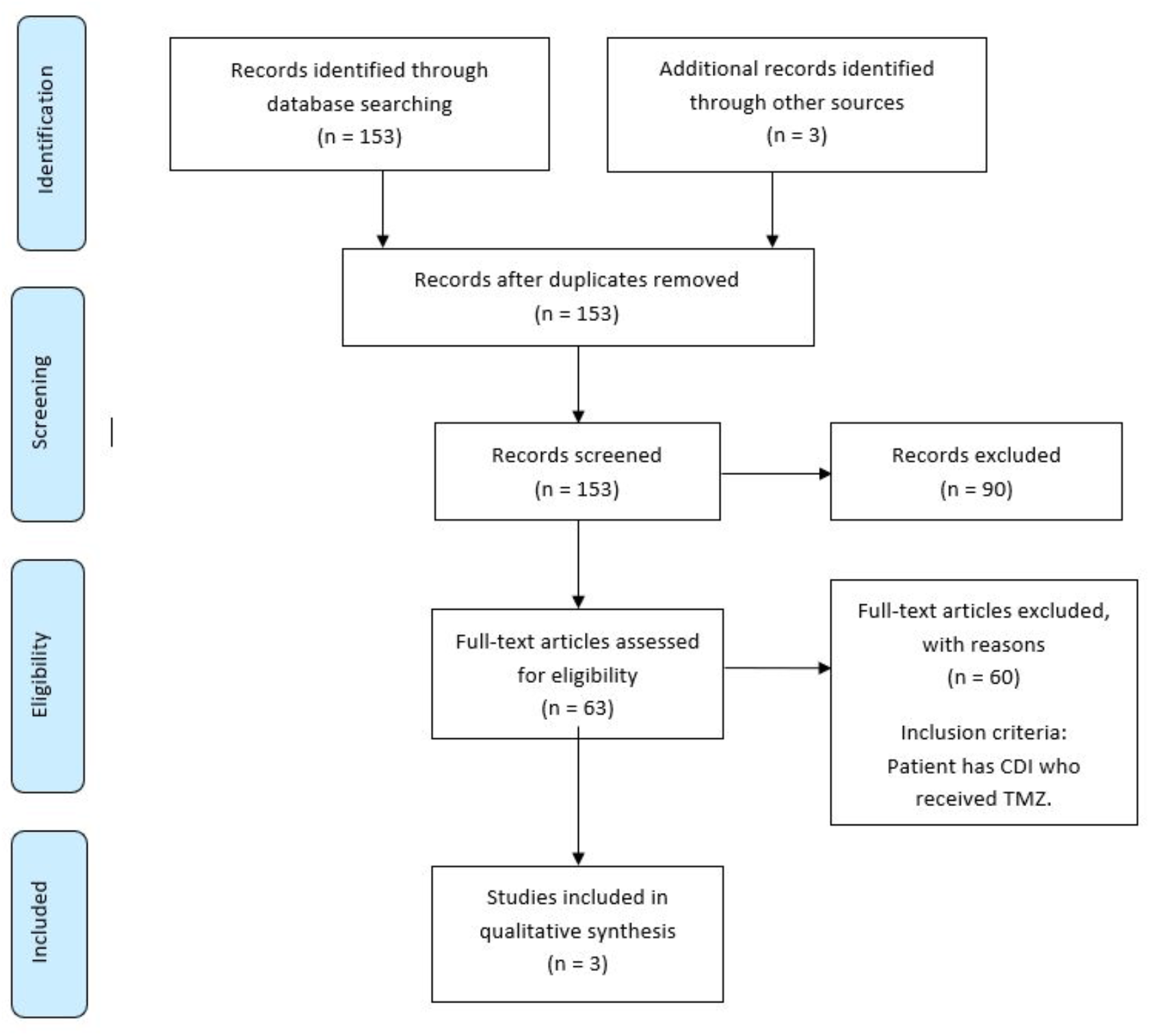
PRISMA flow diagram illustrating the selection process for the studies included in the review for final analysis.

**Figure 2:**
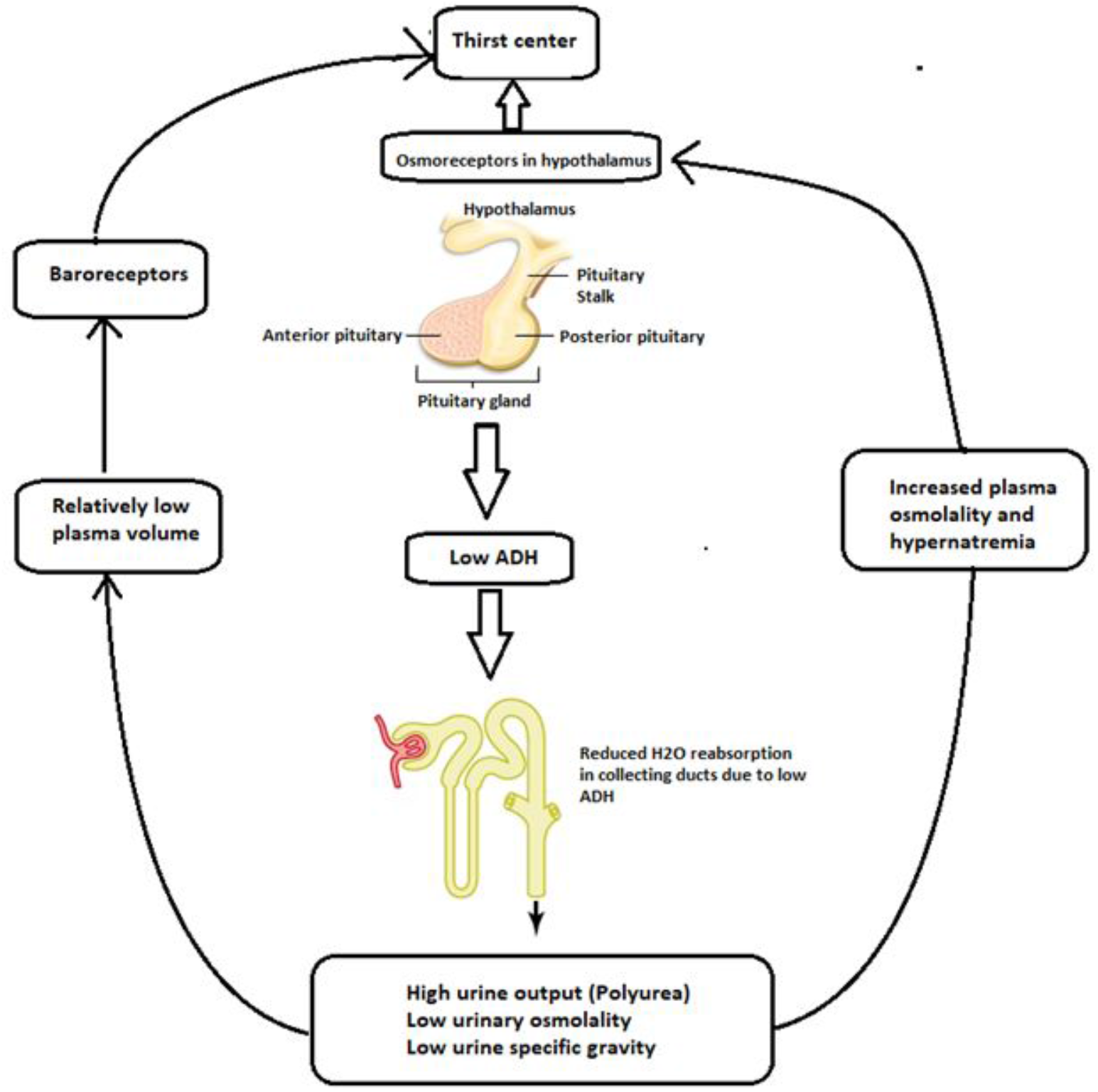
Pathogenesis of CDI and its clinical manifestaions

**Figure 3:**
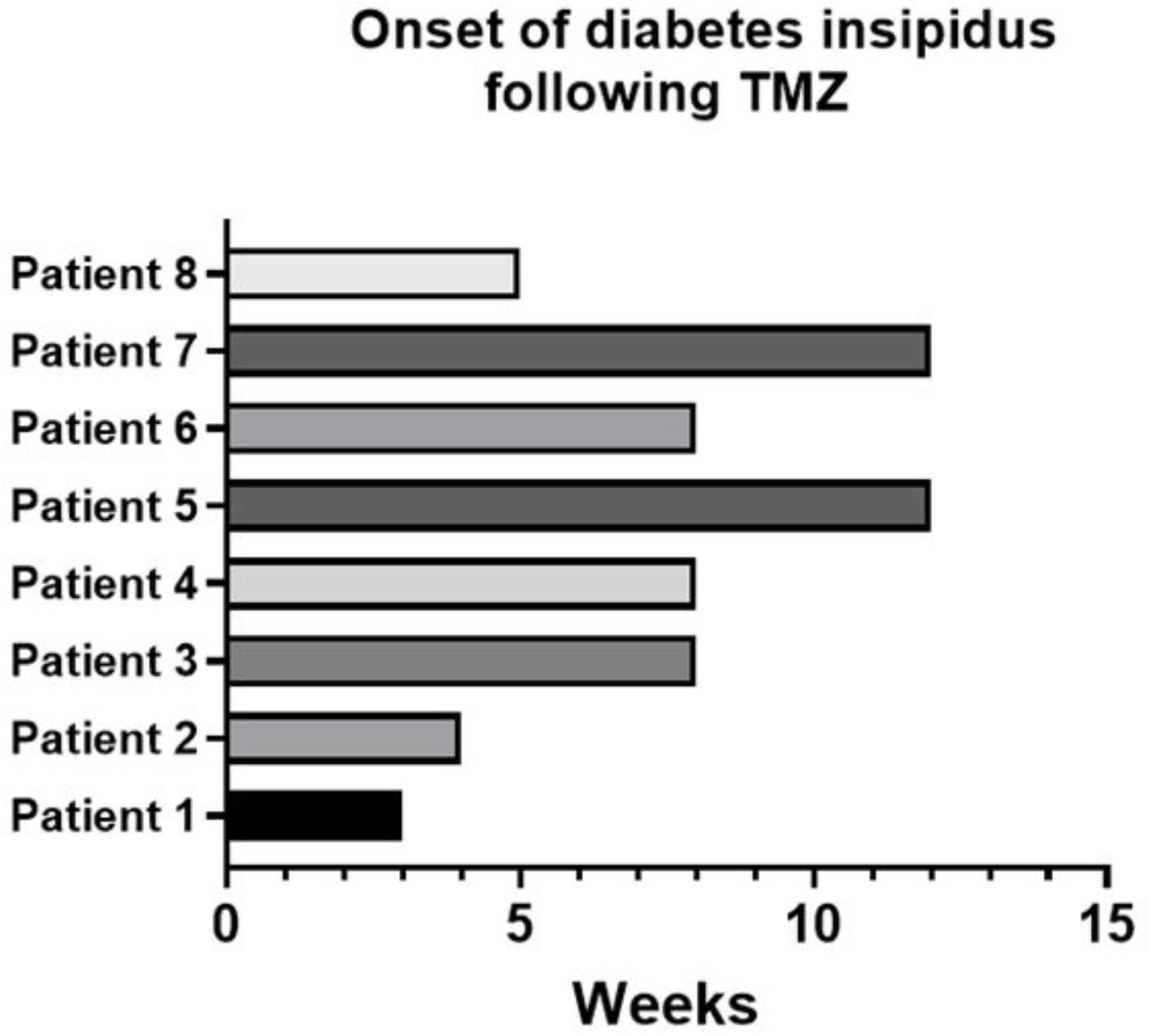
Demonstrating the timeline of developing diabetes insipidus following temozolomide treatment

Temozolomide is an oral alkylating chemotherapeutic agent with a hundred percent oral bioavailability. It is most frequently used for malignant glioma and has been used to treat various malignant CNS tumors, low-grade gliomas, melanoma, and other solid tumors. Temozolomide is a prodrug that spontaneously degrades to its cytotoxic derivative, monomethyl 5-triazeno imidazole carboxamide. This active compound then methylates the purine bases, O6 and N7 of guanine, and N3 position of adenine. These methylated purine bases lead to a continuous cycle of DNA mismatch repair (MMR), resulting in strand breaks, ultimately cellular apoptosis(5). Myelosuppression is the most severe dose-limiting adverse effect. While fatigue, nausea, vomiting, and headache(5). Interestingly, central diabetes insipidus has recently reported a rare side effect and an estimated prevalence of 0.3%(6,7). However, we postulate an underestimation as a symptom of increased urinary frequency was previously reported in TMZ treated patients. It is hypothesized that TMZ may directly affect the ADH synthesis, transport, storage, and/or secretion from the hypothalamus. Unlike CDI secondary to cranial radiation therapy, it is reversible when encountered in TMZ treated patients(8). Besides, although transient and reversible we don’t yet know how promptly we should treat this condition. If we treat it conservatively, it may leave the brain parenchyma in a hyperosmolar environment. We don’t know how it may affect the tumor regression or recurrence.

### Literature Search

A thorough search of several databases from 1990 to October 15, 2020, was limited to ‘English’ and human study. The databases included in the search were Epub Ahead of Print and Ovid MEDLINE(R), In-Process & Other Non-Indexed Citations, and Daily, Ovid Cochrane Central Register of Controlled Trials, Ovid Embase, Ovid Cochrane Database of Systematic Reviews, Web of Science, CINAHL, and Scopus. The search method was developed and applied by an experienced librarian with input from the study’s principal investigator (MR). Used keywords for literature include, ‘central DI’ or ‘Central Diabetes Insipidus’ or ‘CDI’ or ‘Diabetes Insipidus’ AND ‘TMZ’ or ‘temozolomide’ or ‘temodar.’ No restrictions were applied on the patient’s age, gender, ethnicity, or geographical location.

### Screening and data extraction

Abstract and titles of the articles obtained through the literature search were screened for relevance and were performed by two independent authors. Articles were retained in the final list, those met inclusion criteria based on full-text text review. Implemented inclusion criteria were, full paper in English, original case reports or clinical study primarily of any patient with diabetes insipidus or hypernatremia who received TMZ for CNS malignancies, clinical presentation details, and adequate management and follow-up data. Papers were excluded if the clinical picture was unrelated to CDI, review articles, the uncertainty of diagnosis, diabetes insipidus unrelated to TMZ therapy, and conference paper. Full manuscripts of finally selected papers were then obtained, and inclusion finalized. Due to the rarity of reported cases we also included the ‘letter to editors’ in our screening when relevant and met the patient criteria with adequate follow-up data. Afterward, two independent authors extracted data on patient demographics, indications of TMZ treatment, clinical manifestations, onset of symptoms following TMZ treatment, evidence of other hormonal disturbance, medication history, comorbidity, response to desmopressin, follow-up. All data was recorded into a pre-formulated electronic database. A third senior reviewer resolved discrepancies found between the two author’s inputs during extraction.

## Result

A PRISMA flow-diagram was assembled. Using the PRISMA flow diagram, two articles and one letter to the editor with a total of 8 cases were included for final analysis; 7 were adult and 1 pediatric. The mean age for adult cases was 45.86 years (range 38-54 years), and the age of the pediatric case was 12-year. Although all of 8 cases (100%) of evident temozolomide induced central diabetes insipidus occurred in males, the population size is too small to conclude the gender-specificity for this condition. Temozolomide was indicated for various primary and metastatic CNS malignancies (Glioblastoma=2, oligoastrocytoma=1, oligodendroglioma=2, Astrocytoma with Gliomatosis cerebri=1, anaplastic astroblastoma=1, Metastatic Pancreatic NET=1, table 1). No radiological evidence of pituitary or hypothalamic involvement by tumor was demonstrated. All patients (100%) initially presented with polyuria and polydipsia as a manifestation of urine concentrating defect appeared at 3–12 weeks following temozolomide initiation. Serum osmolarity has been reported in 6 patients (ranging from 292-319mOsm/Kg). Serum sodium concentration was reported in 7 patients and has revealed mild hypernatremia (143-151mEq/L) with corresponding urine concentrating defects, documented by the presence of paradoxical urinary hyperosmolarity or low specific gravity.

**Table 1:** patient demographics, presenting symptoms and indication for TMZ

| No. | Author | Patient | Indication for TMZ | Site of the tumor (radiation field) | Symptom onset following TMZ initiation (weeks) | Presenting symptom |
| --- | --- | --- | --- | --- | --- | --- |
| 1 | Mahiat *et al.* | 38, M | GBM | Temporo-occipital (Right) | 3 | Polyuria, Polydipsia, nausea, inappetence, weight loss |
| 2 |  | 54, M | GBM | Frontotemporal (Right) | 4 | Polyuria, Polydipsia, generalized weakness |
| 3 | Faje *et al.* | 53, M | Oligoastrocytoma | Posterior parietal (Right) | 8 | Polydipsia, Polyuria, Nocturia |
| 4 |  | 38, M | Oligodendroglioma | Frontal Lobe (Left) | 8 | Polyuria, Polydipsia, Nocturia |
| 5 |  | 45, M | Oligodendroglioma | Frontal Lobe (Right) | 12 | Polyuria, Polydipsia |
| 6 |  | 49, M | Metastatic Pancreatic NET | Pancreas with mets to liver | 8 | Polyuria, Polydipsia |
| 7 |  | 44, M | Astrocytoma with gliomatosis cerebri | - | 12 | new onset sleeping difficulties, nocturia |
| 8 | Kuo *et al.* | 12, M | Anaplastic astroblastoma | Frontal lobe (left) | 5 | polyuria and polydipsia |

GBM= Glioblastoma multiforme, NET= Neuroendocrine tumors, TMZ= Temozolomide

**Table 2:** Biochemical features

| No. | Author | Patient | Serum osmolality (mOsm/Kg) | Serum Na level (mEq/L) | Urine osmolality (mOsm/kg) | Urine specific gravity | Evidence of pituitary or hypothalamus lesion | other medication | Presence of other endocrine or electrolyte dysfunction |
| --- | --- | --- | --- | --- | --- | --- | --- | --- | --- |
| 1 | Mahiat *et al.* | 38, M | 295 | 145 | 172 | 1.005 | No | No | No |
| 2 |  | 54, M | 296 | 145 | 230 | 1.006 | No | No | No |
| 3 | Faje *et al.* | 53, M | 319 | 151 | NA | 1.005 | No | No | No |
| 4 |  | 38, M | 302 | 143 | 196 | 1.006 | No | No | No |
| 5 |  | 45, M | NA | NA | NA | NA | No | No | NA |
| 6 |  | 49, M | NA | 143-146 | NA | 1.003<br>-1.006 | No | TMP-SMX | NA |
| 7 |  | 44, M | NA | 142-145 | NA | 1.006<br>-1.010 | No | Bevacizumab | NA |
| 8 | Kuo *et al.* | 12, M | 305 | 149 | 194 | 1.005 | No | No | No |

TMP-SMX= Trimethoprim-sulfamethoxazole

**Table 3:**
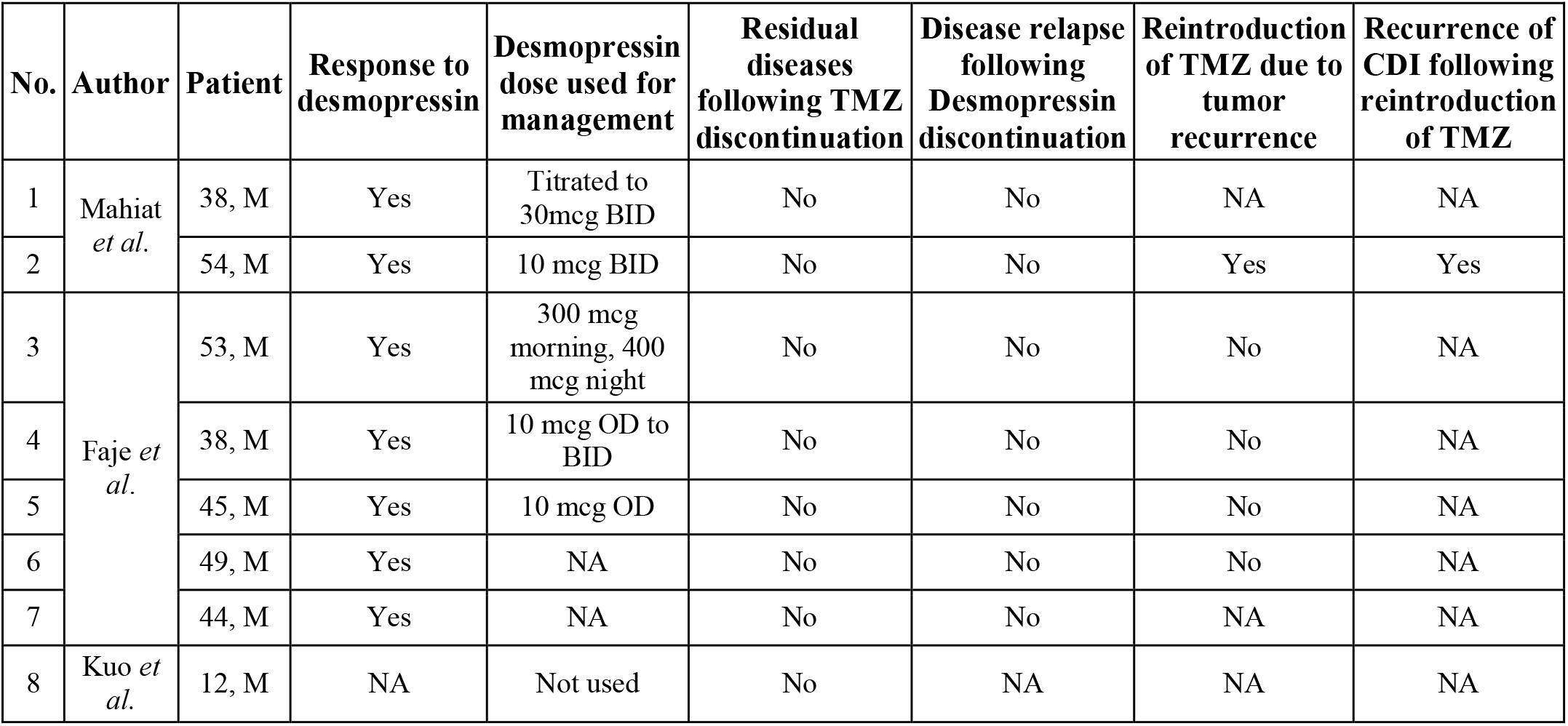
Management and outcome data

## Discussion

Polyuria-polydipsia syndrome may have underlying primary polydipsia, osmotic diuresis, central and nephrogenic DI. Among these, osmotic diuresis demonstrated by high urine osmolality or specific gravity is seen along with hyperglycemia, resulting from steroids used for a tumor or therapy-associated cerebral edema (1,9). Whereas diabetes insipidus is characterized by hypo-osmolar and low specific gravity urine despite hyperosmolar and hypernatremic plasma. In contrast to diabetes insipidus, patients with primary psychogenic polydipsia exhibit hyponatremia instead of hypernatremia. All reported cases in this review were adiabatic that made the possibility of osmotic diuresis unlikely, and all revealed low urine osmolality, specific gravity, and high serum sodium concentration.

Serum and urinary osmolarity testing following overnight water deprivation confirm the defective urine concentrating capacity, thus diabetes insipidus, which further differentiated by evaluating urine osmolarity following exogenous desmopressin (dDAVP) administrations. However, the complete reversal of polyuria-polydipsia syndrome, along with the ascent of urine osmolality and the fall in serum sodium level and plasma osmolality following dDAVP, strongly suggest the presence of central DI(1,9,10). Although a water deprivation test is considered for the gold standard to diagnose diabetes insipidus, it was not reported in any of the cases. However, all patients improved symptomatically, and hypernatremia was resolved following the desmopressin trial, which indicates the presence of ADH deficiency(10). All reported clinical and laboratory abnormalities suggestive of central DI were well correlated with initiation and discontinuation of TMZ and desmopressin therapy. Furthermore, Mahiat *et al*. reported a case that demonstrated a recurrence of the similar clinical and laboratory picture on the TMZ rechallenge test with lower dosing strategy and correction clinical and laboratory parameters following re-administration of dDAVP(7). Laboratory testing of hypothalamic-pituitary hormones was reported on six patients and was in the physiological range. We hypothesize, intact thirst and relatively mild nature as you mentioned are responsible for borderline lab values

The current standard regimen with temozolomide for malignant glioma begins with focal radiation over six weeks with concomitant daily TMZ, after that six cycles of adjuvant therapy for five days for every 28 days. However, more intensive dosing paradigms have also been reported(11). Five patients, including the pediatric case, received daily TMZ, one patient was treated with daily dosing on alternate weeks.

However, it is inconclusive whether TMZ-induced diabetes insipidus is dose-dependent or not. Admittedly, radiation therapy may lead to hypothalamic/pituitary deficit. Although anterior pituitary hypofunction has reported following therapeutic range cranial radiation for the extrasellar brain tumors, DI has never been reported. Moreover, radiation-induced neuroendocrine hypofunction is supposed to be irreversible. All of the reported cases revealed complete remission following temozolomide cessation. A very few TMZ-induced CDI cases have been reported despite its wide use for various brain tumors since accelerated approval by the US FDA (Food and Drug Administration) 1999(11–13). Faje *et al*. found that the prevalence of diabetes insipidus in TMZ treated patients was 0.3% on their electronic data-based survey 1545 patients received TMZ treatment (6,14). Authors also identified 6-8% of patients who reported urinary incontinence and increased urinary frequency; however, it was not possible to reveal a urinary concentrating defect in these patients. Mahiat *et al*. prevalence of 0.28% on their survey using pharmacy electronic data, is very similar to that documented by Faje et al. (6,7,15)

Athavale *et al*. reported a case of demonstrating acute interstitial nephritis (AIN) along with nephrogenic diabetes insipidus following temozolomide and sulfamethoxazole-trimethoprim treatment (16). Renal biopsy revealed focal tubulointerstitial change and interstitial inflammation. The patient’s renal function was improved by discontinuing sulfamethoxazole-trimethoprim and treatment with thiazide diuretics for nephrogenic DI without further deterioration with resuming temozolomide. This concludes TMZ was not the culprit drug for nephrogenic DI.

The pathophysiology of central DI secondary to TMZ is unclear. Several hypotheses have been proposed including, impaired synthesis, storage, or secretion from the hypothalamus of the neurohypophysis. Proposed mechanisms are cytoskeletal disarray of neuroendocrine cells, disrupted receptor signaling by TMZ in magnocellular neurons responsible for ADH synthesis and secretion. Interestingly, ADH deficiency has never been reported in association with a prodrug of TMZ, dacarbazine, having poor blood-brain barrier penetration (7,17). Another possibility remains that the adverse effect is an idiosyncratic drug reaction to genetically susceptible individuals. Just like most idiosyncratic reactions the occurrence of CDI in our included studies was very rare. In both of our included studies, the prevalence was described as 0.28% and 0.32% respectively among patients who received TMZ therapy. Idiosyncratic reaction is specific to individuals & this was demonstrated in one of our study patients (#2) by the recurrence of symptoms following a symptom-free interval only after reintroduction of TMZ therapy. A characteristic of an idiosyncratic effect is that it’s dose independent. In our study, the patient who had a recurrence of symptoms following the reintroduction of TMZ did so in two different dosing regimens. The first episode was the initial, more intense daily TMZ therapy concomitant with radiotherapy. While the second episode was after a rechallenge of TMZ on a less intensive dosing strategy (150 mg/m2 for 5 days in 28 days cycle). Many but not all idiosyncratic reactions are seen in genetically predisposed individuals (18). For example, abacavir hypersensitivity reactions in HLA-B*57:01 patients. We hypothesize that the genome responsible for central DI following TMZ may be located in the Y chromosome. This argument further benefits from the fact that all of our study patients were male. Overall, the symptoms and lab finding in our patients suggest the central diabetes insipidus / polyuria-polydipsia syndrome resulting as an adverse reaction to Temozolomide is relatively mild in nature and reversible with rapid response to DDAVP in all patients; however their effect on morbidity and overall prognosis of cancer patients remains to be discussed. some probabilities remain that DI symptoms were induced by brain surgery. 5 out of 8 patients in our study cohort did undergo surgery before initiation of TMZ. We hypothesize the symptoms are due to TMZ as symptoms started after initiation of TMZ therapy in all patients as well as the fact that patient-6 did not undergo any type of brain manipulation. And patient-4 had his last craniotomy done 2 years before TMZ initiation and symptoms began only after 8 weeks of therapy. Besides, reproducible symptoms with restarting TMZ make the phenomenon scientifically believable.

As like other this review is not be beyond limitations. There was insufficient data on maintenance TMZ dosing and cycle length for 3 to 7. While most endocrinologists would include urine output > 250cc/h for three consecutive hours to diagnose CDI, urine output data are missing from the reported cases. Further study and watchful monitoring of patients is required to detect clinical relevance and robustness of speculated pathophysiology.

## Conclusion

Central DI appears to be a scarce adverse effect of TMZ. We postulate that the current prevalence might be underestimated due to a lack of clinical suspicion and proper evaluation of any patient with polyuria polydipsia symptom on TMZ regiment. All patients with TMZ therapy should be aware of CDI symptoms before starting treatment with instructions on the expected urinary frequency and monitoring of intake, output, and thirst. The emergence of any concerning symptoms should thoroughly be evaluated to detect any possibility of ADH deficiency. Although mild to moderate hypernatremia due to CDI can be managed conservatively, future treatment protocol regarding this scenario should be considered. This review encourages the necessity of rapid detection of this side effect where prompt management can remarkably improve the patient’s quality of life while on TMZ therapy.

## Data Availability

Data are available in tables and diagrams.

## Human/Animal Rights

This article does not contain any studies with human or animal subjects performed by any of the authors.

## Acknowledgments

We want to acknowledge and thank Leslie Hassett, Outreach Librarian, Mayo Clinic Libraries, who helped in the comprehensive literature search and CMSR (center for medical study and research) foundation.

